# Characterisation, identification, clustering, and classification of disease

**DOI:** 10.1101/2020.11.26.20227629

**Authors:** A.J. Webster, K. Gaitskell, I. Turnbull, B.J. Cairns, R. Clarke

## Abstract

Data-driven classifications are improving statistical power and refining prognoses for a range of respiratory, infectious, autoimmune, and neurological diseases. Studies have used molecular information, age of disease incidence, and sequences of disease onset (“disease trajectories”). Here we consider whether easily measured risk factors such as height and BMI can usefully characterise diseases in UK Biobank data, combining established statistical methods in new but rigorous ways to provide clinically relevant comparisons and clusters of disease. Over 400 common diseases were selected for study on the basis of clinical and epidemiological criteria, and a conventional proportional hazards model was used to estimate associations with 12 established risk factors. Comparing men and women, several diseases had strongly sex-dependent associations of disease risk with BMI. Despite this, a large proportion of diseases affecting both sexes could be identified by their risk factors, and equivalent diseases tended to cluster adjacently. This included 10 diseases presently classified as “Symptoms, signs, and abnormal clinical and laboratory findings, not elsewhere classified”. Many clusters are associated with a shared, known pathogenesis, others suggest likely but presently unconfirmed causes. The specificity of associations and shared pathogenesis of many clustered diseases, provide a new perspective on the interactions between biological pathways, risk factors, and patterns of disease such as multimorbidity.

---

John Graunt’s pioneering epidemiological studies in the 1600s required the identification and clustering of symptoms into disease types with similar aetiologies ^1^. Clusters needed to be fine enough to distinguish different underlying causes, but coarse enough to allow meaningful statistical study. The modern International Classification of Diseases (ICD) ^2,3^ assigns each disease a hierarchical code in which successive digits provide increasing detail about the cause, pathology, or anatomical site of the disease, and it continues to evolve ^4^.

Data-driven classification of disease is a recent idea, made possible by access to large population studies, such as UK Biobank ^5^. Examples include using molecular or imaging data to identify and classify subtypes of disease such as metabolic syndrome ^6^, amyotrophic lateral sclerosis (ALS) ^7^, cancer ^8,9^, arthritis ^10,11^, and dengue fever severity ^12^. The subtypes allow more accurate prognoses for disease severity ^12^, co-morbidities ^13^, outcomes ^7,14^, and response to treatment ^15^. More general studies intend to better characterise the phenotype ^6,9,13,16–26^, using either molecular or genomic data ^6,9,13,17–19^, or the times and sequences of disease incidence (“disease trajectories”) ^20–26^. Aims include improved aetiological understanding ^13,16,19,27^, quicker and more accurate diagnoses ^15–17^, more detailed prognoses ^15–17,20,21,23,24^, improved statistical power ^28^, improved care ^19,20,22^, and facilitating drug development ^15,29^. Improved classification schemes are widely expected to improve our understanding of disease ^7–13,16,27^ and the precision of drug targets and clinical trial design ^15,28,29^, accelerating advances such as personalised medicine ^29^ and improving our ability to understand, and prevent, both individual and multiple diseases including multi-morbidity ^30^.

Previous data-driven classifications have considered molecular data, the time of disease onset, or the sequences of diagnosed diseases (“disease trajectories”). Here we explore whether easily measured, well-known risk factors such as height and body mass index can be used to usefully characterise, identify, and cluster diseases.

## Results

Data are from the UK Biobank cohort of over 500,000 men and women aged between 40 and 69 years, recruited during 2006-2010. For inclusion, diseases were the primary clinical diagnosis recorded in hospital records with an ICD-10 code between 31 March 1996 and 31 March 2017, that refer to a clear diagnosis of a health-related disease. Diseases were selected by a statistician and two clinical epidemiologists, on the basis of statistical and clinical criteria, as detailed in the Methods and summarised in table 1.

**Table 1:** Selection criteria for clustering of diseases

| Clinical inclusion criteria |  | Eligible hospital episodes |  |
| --- | --- | --- | --- |
| Prior disease | Diseases were the first primary hospital diagnosis in each ICD-10 chapter | 425383 | male |
|  |  | 502771 | female |
| + Clinical considerations | Clinically distinct, age-related disease, or R-coded diseases of unknown aetiology | 400006 | male |
|  |  | 468398 | female |
| Statistical inclusion criteria |  | Eligible diseases |  |
| Successful fit | At least 50 cases, and a covariance matrix with no unusually large or small eigen-values that exceed the mean by 2.5 standard deviations when outliers were included | 343 | male |
|  |  | 346 | female |
| + Statistically significant | Statistically significant risk factors at the 0.05 level, after a multiple-testing Bonferroni adjusted multivariate $\chi^2$ test | 150 | male |
|  |  | 140 | female |
| + Proportional hazards | Test of proportional hazards assumption - no statistically significant deviation at the 0.05 level, after a multiple-testing FDR adjustment | 138 | male |
|  |  | 127 | female |
| + Unisex | The set must include the same disease in both men and women | 86 | male and female |

All results describe diagnoses that were an individual’s first primary diagnosis in an ICD-10 chapter. This compromise between reducing the risk of confounding by prior disease and retaining sufficient cases was later tested by a sensitivity analysis. We considered the well-known risk factors of: diabetes, height, body mass index (BMI), smoking status, systolic blood pressure (SBP), alcohol consumption, and walking pace, and adjusted for the established confounders and female-specific risk factors of: deprivation tertile, education, hormone replacement therapy (HRT) (women only), and having one or more children (women only). As detailed in the Methods, risk-factor associations were estimated using a proportional hazards model using age as the time variable, left-truncated at the study start and right-censored at the study end or any cancer other than non-melanoma skin cancer. To reduce confounding by age we stratified by year of birth (YOB), and adjusted by the age at which participants joined the study. To maximise the number of cases in each category we assumed a linear response to the continuous measures of BMI, height, and SBP. Non-linear associations would reduce the accuracy of fits, and leads us to argue against inferring causal associations between risk factors. We used well-known and biologically meaningful variables to aid interpretation of disease clusters, but as measurable recognised physical characteristics, it would be acceptable if “risk factors” were symptoms.

Figure 1 shows the number of diseases with statistically significant risk factors, that increase with the number of cases due to maximum likelihood estimates becoming increasingly accurate and identifying smaller effect sizes. Overall there were smaller proportions of statistically significant associations with injuries or symptoms of unknown origin. There were similar numbers of chronic and acute diseases with 230 or more cases, but rarer diseases with 49-230 cases were almost twice as likely to be acute than chronic disease. Despite infectious diseases needing exposure to an infectious agent to trigger an infection, there were clear associations of infectious diseases with risk factors, possibly because we have selected infectious diseases that are likely to reflect an underlying susceptibility. The proportion of diseases failing a proportional hazards test increased with the number of cases, presumably because the test became more sensitive. With larger datasets, it is possible that fewer diseases will satisfy the proportional hazards model, although the failure might be correctable with a different combination or increased number of risk factors.

**Figure 1:**
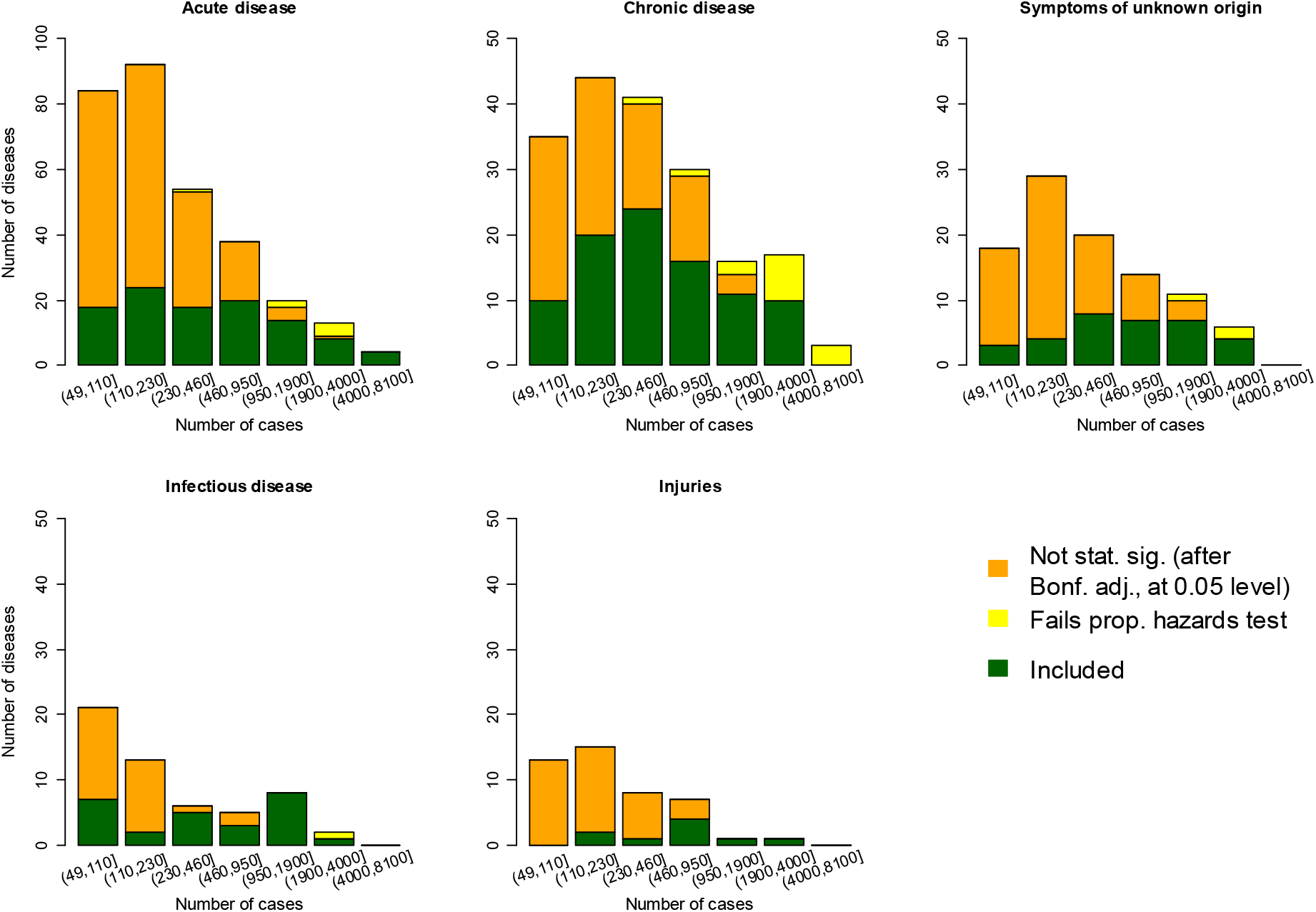
689 diseases of men or women were categorised as acute, chronic, infectious, injuries, or symptoms of unknown cause (separate plots), and grouped by the number of cases (horizontal axes). We considered: Are associations statistically significant at the 0.05 level after a Bonferroni multiple-testing adjustment? - no (orange), yes: Are tests consistent with a proportional hazards model? - no (yellow), yes (green). The median number of cases was 214. The vertical axis for acute diseases has a different scale.

To compare diseases, we were interested in strong biologically meaningful comparisons, for example between current smokers and a baseline of never smokers, as opposed to a baseline of previous smokers. Such substantial differences are more likely to be associated with changes to biological pathways that can modify disease risk. Because the maximum likelihood estimates (MLEs) for parameters are normally distributed, the distribution for a subset of parameters is easily obtained by marginalisation ^31^. The mean and covariance matrices of a subset are simply the rows and columns of the mean and covariance matrices that correspond to the parameters of interest ^31^. These values are generally quite different than those obtained by fitting the subset of parameters directly. This allowed us to adjust for parameters that are known to influence disease risk, but for clustering and comparison we used marginalisation to solely consider: BMI, height, SBP, slow walking pace (versus fast walking pace), regular drinker (versus rarely drink), and current smoker (versus never smokers). The procedure also ensures that each risk factor is represented by a single variable when clustering, reducing the potential for clustering to be dominated by a single risk factor (e.g. a categorical variable with *d* levels would otherwise be represented by *d* parameters when clustering).

### Identification of disease

Each disease present in both men and women were assigned to the one with minimum Battacharyya distance between their estimated associations with potential risk factors. The proportion of diseases matched to their equivalent disease in the opposite sex are plotted in figure 2, grouped as acute, chronic, infectious diseases, and symptoms of unknown origin (R-codes). For 38% of the 172 diseases considered, the nearest disease measured by Battacharyya distance was the equivalent disease in the opposite sex, and for 80% of diseases the equivalent disease was among the nearest 8 diseases (the nearest 5%).

**Figure 2:**
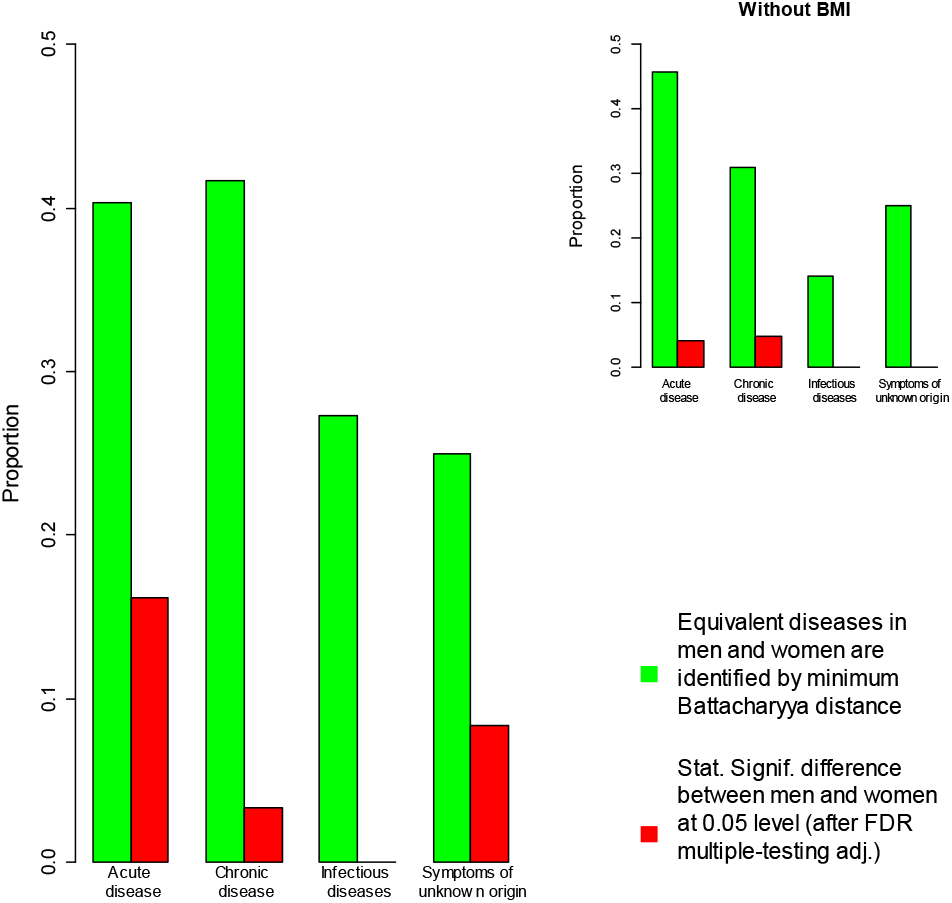
The proportion of diseases whose equivalent disease in the opposite sex has the smallest Battacharyya distance is plotted in green. The proportion of diseases with statistically significant differences between men and women are plotted in red. The differences are mainly due to different associations with BMI (inset).

### Differences between men and women

The proportions of diseases with statistically significant differences in their associations with risk factors are shown in figure 2. Approximately 5% of diseases had statistically significant differences between men and women at the 0.05 level after an FDR multiple-testing adjustment ^32^, and this dropped to ∼1% when BMI was excluded as a risk factor.

The risk factors responsible for statistically significant differences between men and women are considered in figure 3. The heat map indicates whether a risk-factor is associated with a higher risk for women (red), or lower risk (white), with orange neutral. Because BMI appeared to have different risk associations in men and women, it was removed and the analysis rerun. Removing BMI reduced the number of diseases with statistically significant risk factors (after a Bonferroni adjustment), from 172 to 156. Figure 2 shows that the proportion of diseases with statistically significant differences between men and women reduced from *∼* 5% to *∼* 1%, and figure 3 shows that those diseases are arthrosis of the knee and kidney stones. The differences do not appear to be solely due to any particular risk factor. A sensitivity analysis with sex-dependent tertiles replacing continuous measurements, found similar results (see Supporting Information, figure 3). Overall we found strong evidence for sex-specific associations in some diseases affecting men and women, especially for BMI.

**Figure 3:**
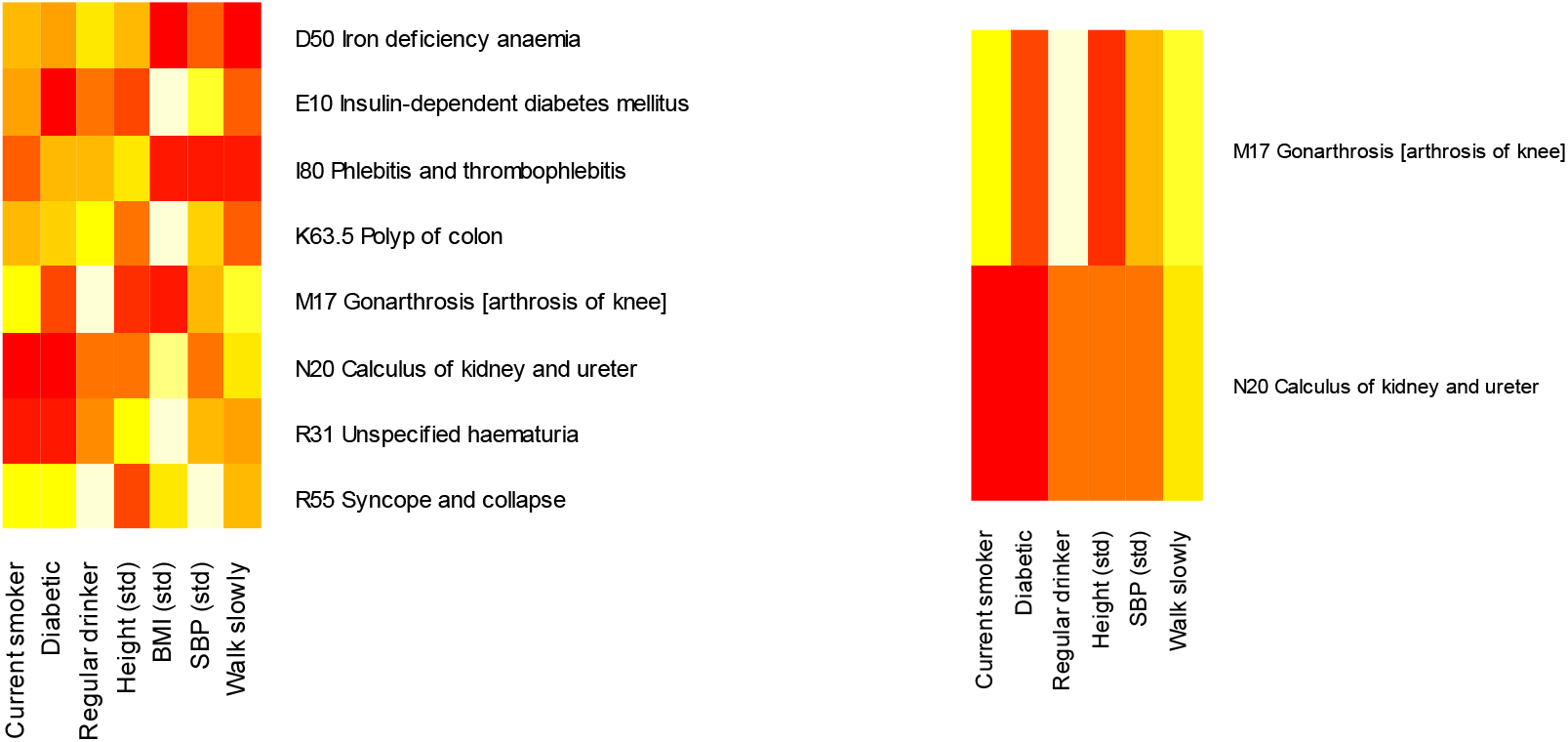
Disease pairs with statistically significant differences in their associations with risk factors at the 0.05 level after an FDR multiple-testing adjustment. With all associations (left), and without BMI (right). Red indicates an association with higher risk for women than men, white a lower risk, and orange neutral. Without BMI as a risk factor, only two diseases continue to have statistically significant differences.

### Clustering of disease

Figure 2 shows that many diseases could be identified by their associations with well-known risk factors. Presuming the associations reflect common aetiological pathways, then clustering by them may yield clusters of diseases with similar aetiologies. Hierarchical clustering was used to capture and visualise similarities between the risk factors for disease, and generated a hierarchical structure of increasingly similar clusters. The dendrogram is coloured to indicate 24 groups. The clustering is shown in figure 4, along with a heat map for the risk factors associated with each disease. This allows us to simultaneously visualise how diseases cluster, and the associations responsible for the clusterings.

**Figure 4:**
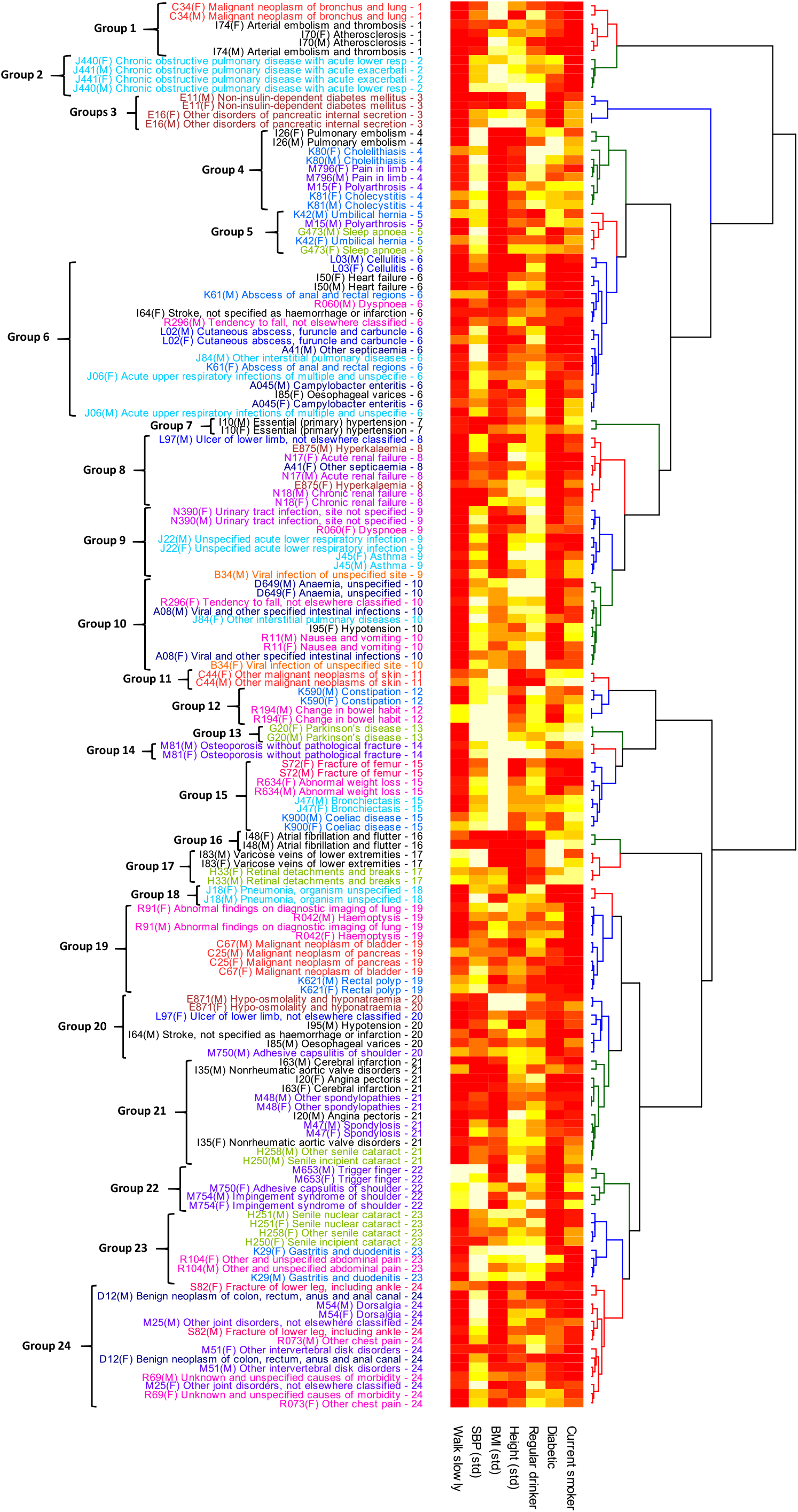
The estimated fitting parameters and their covariance matrices were used to calculate the Battacharyya distances between diseases, and clustered hierarchically using the Ward.D2 algorithm. Diseases in men and women tend to cluster adjacently. Labels are coloured by their first ICD-10 digit, and the dendrogram is coloured with the top 24 groups in the cluster (see figure 5). Associations with potential risk factors are indicated by the heat map, with red an association with higher risk, white with lower risk, and orange neutral.

When considering figure 4 it is useful to note that: (1) Disease descriptions with the same first-digit of ICD-10 code are coloured the same, e.g. I50 and I70 are both coloured black. (2) If the same disease in men and women cluster together, then it is likely to have a distinctive combination and magnitude of associations with risk factors. (3) Any diseases connected by a tree with small depth will have a quantitatively similar combination of associations. (4) The heatmap indicates a cluster’s association with risk factors, with red associated with higher risk, white with lower risk, and orange neutral. For example, considering figure 4, chronic obstructive pulmonary disease, lung cancer, arterial embolism, and atherosclerosis are clustered closely together (groups 1 and 2), and are being identified primarily by the increased risk associated with smoking and walking slowly, with the magnitude of associations producing the finer subgrouping.

### Symptoms, signs, and abnormal clinical and laboratory findings, not elsewhere classified

Chapter XVIII of ICD-10 is devoted to “Symptoms, signs, and abnormal clinical and laboratory findings, not elsewhere classified” ^2,3^, and accounted for 11% of primary hospital episodes in the UK Biobank data. Despite their uncertain aetiology, 60 of the 98 diseases in men or women had statistically significant risk factors at the 0.05 level after an FDR multiple-testing adjustment, and 36 were statistically significant at the 0.05 level after a Bonferroni adjustment. Ten diseases that satisfied the FDR-adjusted proportional hazards test and were also present in both men and women were included in the clustering studies, and for most of these their risk associations were similar in men and women (figure 4, R-coded disease descriptions).

### Confounding by prior disease

Because the same individual’s data can appear every time a hospital episode has a primary disease from a different ICD-10 chapter, there is potential for confounding by prior disease. To test whether this influenced the clustering results, we took the 24 clusters in figure 4 and refit the proportional hazards model for each disease, but now excluded data with any prior diseases from the same cluster as the disease being studied. This prevented the clustering of diseases from different chapters being influenced by repeat hospital episodes from the same individuals.

Despite having fewer cases, the resulting cluster is almost identical to figure 4 (see Supporting Information, figure 2), with all pairs of clustered diseases continuing to cluster with each other. This strongly suggests that the clusters were driven by similarities in risk factors as intended, not by sequences of prior diseases.

### Tertiles versus a continuous linear model

By assuming a proportional hazards model with a simple linear relationship between continuous measurements for height, BMI, and SBP, it was possible to consider diseases with fewer cases than needed by a more complex model. To test the sensitivity of clusterings to this linear approximation, we refit a proportional hazards model to the same set of diseases but with sex-specific tertiles for height, BMI, SBP, and year of birth. Before clustering we again used marginalisation to compare a baseline of non-smokers, non-diabetic, rarely drink, and minimum tertiles for height, BMI, and SBP, to parameters for regularly smoking, diabetes, regularly drinking, and maximum tertiles for height, BMI, and SBP. We did not require fits to satisfy any statistical tests because the fewer numbers of cases in each tertile were expected to make the fits poor for some diseases.

As shown in the Supporting Information figure 3, the resulting clusters are similar, with 54% of all pairs of clustered diseases remaining together after reanalysing with tertiles. The disease of arterial embolism and thrombosis (I74) was missing from the analysis because there were too few cases in women when the analysis used tertiles. Ten diseases were most sensitive to the model being fit, having no other disease clustered with them in both clusterings. These were: H25.8 - Other senile cataract (men), K42 - Umbilical hernia (men), K59 - Constipation (men), K61 - Abscess of anal and rectal regions (men), L97 - Ulcer of lower limb (women), M15 - Polyarthrosis (men), M51 - Other intervertebral disk disorders (women), R11 - Nausea and vomiting (men), R29.6 - Tendency to fall (men), R69 - Undetermined causes of morbidity (men).

Diseases with statistically significant differences between men and women were also similar (see Supporting Information, figure 4). The differing analyses found 4 diseases common to both studies with statistically significant differences at the 0.05 level after an FDR multiple-testing adjustment ^32^. Without BMI as a risk factor, both studies found that kidney stones (N20) continued to have different risk associations for men and women.

## Discussion

The broad systematic study of sex-specific diseases, specificity of observed associations, and shared pathogenesis of many clustered diseases offers potential new insights into the clinical presentation and aetiopathology of disease, some of which are explored below.

### Sex differences and epidemiological practic

There is a growing awareness of differences between men and women for the incidence, diagnosis, prognosis, and treatment of disease ^25,33^. Sex-dependent risk factors have also been found in association with cardiovascular disease ^34^. Here we find a substantial proportion of diseases with different risk associations between men and women, for BMI in particular (figures 2 and 3). Further work is needed to understand the causes and implications of different risk associations, but the sex-dependent differences for BMI in particular, are sufficiently clear that they should be accounted for in future studies.

The proportional hazards model failed more frequently as the number of cases increased (figure 1). For larger data sets in particular, the model should be tested, and modified as required. With sufficient data, alternative methods may need to be considered.

### Specificity of associations

Despite 5% of diseases having substantially different associations between risk factors in men and women, 38% of diseases were correctly identified with their equivalent disease in the opposite sex, and 80% had their equivalent disease among the nearest 8 (of 172) diseases. This would only be possible if men and women had similar quantitative associations with risk factors for a given disease, and if these are sufficiently distinct from those for other diseases. The influence of risk factors on disease onset seems surprisingly specific in many cases, and with more risk factors this specificity may increase. For example, if the 7 risk factors had a trinary value of e.g. tertiles, there would be 3^7^ = 2187 possible combinations, but if the number of risk factors were doubled from 7 to 14 the combinations would exceed 4 million. In principle, it may be possible to define diseases by their response to a specific set of risk factors.

### Pathways for disease

An underlying reason for this work was to explore whether clustering by common risk factors can help identify pathways for disease. For example, renal failure, hyperkalaemia, and ulcers of lower limbs in men are clustered in group 6, along with other septicaemia in women. Renal failure can increase the risk of ulcers of the lower limb ^35,36^, and hyperkalemia can be caused by kidney disease. However, a sensitivity analysis excluded prior diseases from the same cluster prior to fitting the proportional hazards model, and produced an almost identical clustering of diseases, consistent with clusters being driven by associations with risk factors (as intended), not prior disease. One interpretation is that the disease cluster is driven by a common pathway such as atherosclerosis, with some associations being risk factors for it, others symptoms of it, and the diseases a consequence of it. This could produce (non-causal) associations between subsequent hospital admissions for different diseases. In contrast, cardiovascular diseases such as arterial embolism, pulmonary embolism, and atrial fibrillation are from the same ICD-10 chapter, but have different underlying causes, and are found in different clusters with quantitatively different risk associations.

Cardiovascular diseases appear in several different clusters, suggesting they are influenced by a range of different pathways for disease onset or severity. Arterial embolism and atherosclerosis are clustered with lung cancer in group 1, and adjacent to chronic obstructive pulmonary disease (COPD) in group 2, suggesting a similar and possibly smoking-related cause. Pulmonary embolism is in a group of 9 diseases (group 4) that includes gall-stones, pain in limb, and polyarthrosis in women. Gallstones have previously been associated with an higher risk of pulmonary embolism ^37^, that was attenuated after cholecystectomy ^37^. Heart failure and unspecified stroke in women appear in a large group of 18 diseases, in which 11 of the remaining 15 diseases involve infections. Atrial fibrillation has sufficiently specific associations to be clustered on its own in group 16. Non-rheumatic aortic valve disorders, angina pectoris, and cerebral infarction, appear in group 21, along with spondylosis, other spondylopathies, and senile cataracts in men. Cervical spondylosis (CS) have previously been associated with a higher risk of posterior circulation infarcts ^38^, and with acute coronary syndrome ^39^. Unspecified stroke, hypotension, and oesophageal varices, all in men, are in the adjacent group 20, along with hypo-osmolarity and hyponatraemia, and ulcer of lower limbs in women.

The majority of diseases involving infections are in a large cluster (group 6), described above in the context of cardiovascular diseases. The clustering suggests that susceptibility or severity could be mediated by a common underlying pathway. There is nothing unusual about the associations with walking slowly, diabetes, high BMI, and smoking, suggesting that the specific strengths of those associations are producing the cluster. Four other types of infections affecting both men and women (eight diseases), are in groups 9 and 10, and appear to have weaker associations with smoking and BMI than those in group 6.

### Identification and re-classification of disease

Many diseases of uncertain aetiology (R-coded diseases in ICD-10), had statistically significant risk factors, often sufficiently specific for equivalent diseases in men and women to cluster adjacently (figure 4). This could be explained by hospital referrals being influenced by specific risk factors and symptoms, as specified by medical training or guidelines. Alternatively, the quantitative disease-specific patterns of associations between risk factors and R-coded diseases could reflect an underlying pathophysiological cause. From the perspective of the Bradford Hill criteria ^40,41^: *Strength of association, Consistency, Specificity, and Temporality* - there were strong, dose-related, statistically significant, disease-specific, subsequent responses to risk factors in both men and women. *Analogy, Plausibility, and Coherence* - like all diseases, evidence of disease is sufficiently strong and specific for hospital admission and identification with one of nearly 100 R-coded diseases.

R-coded diseases are rarely discussed or studied, so it is worth examining the diseases with which they cluster in detail: (1) Nausea and vomiting is clustered with specified intestinal infections, suggesting a possible infectious origin. The cluster also contains anaemia, and diseases in women-only of tendency to fall, other interstitial pulmonary diseases, hypotension, and viral infections of unspecified site. (2) Change in bowel habit is clustered with constipation in group 12. (3) Abnormal weight loss is clustered with fractures of the femur, bronchiectasis, and coeliac disease in group 15. Weight loss is a potential cause of fractures that are mediated by osteoporosis, but similar risk associations for weight loss and femoral fractures would suggest that weight loss could be a symptom of an unidentified underlying process. (4) Abnormal findings or imaging of lung, and haemoptysis, are clustered with pancreatic and bladder cancers, and rectal polyp, in group 19. (5) Other and unspecified abdominal pain is clustered with gastritis and duodenitis in group 15. We are unaware of any indirect reasons why the risk factors for diseases with such similar symptoms would coincide, but the group also contains four cataract diseases, that seem most likely due to coincidental similarities between the risk associations. (6) Other chest pain and undetermined causes of morbidity are in group 24, a group that also includes back pain, intervertebral disc disorders, other joint disorders not classified elsewhere, fractures of the lower leg, and benign neoplasms of the colon, rectum, and anus. The links between these undiagnosed causes of pain and morbidity, and diagnoses of back and intestinal problems may be relevant for improving the accuracy of diagnoses. (8) A few other R-coded diseases are included, but these diseases appear in different clusters for men and women, and are not discussed further.

## Limitations

Many of the limitations here are inherent to any cohort study, but some are accentuated by the need to simultaneously study multiple diseases. *Disease selection:* Uncertainty about the history of treatment decisions made it impractical to identify and exclude diseases whose hospital episode rates have geographical or temporal variations due to changes in diagnosis or treatment practices, such as a change in reported incidence of sepsis due to changes in coding ^42^. Instead we relied on statistical tests to detect when large variations in episode rates were causing statistical models to fail or lose power. *Cohort:* Due to the minimum age of participants in UK Biobank, we can only study diseases of old age, and the UK Biobank cohort is not representative of the UK or global population. Hospital referrals, diagnoses, and recordings of diagnoses are all biased by clinical procedures and training. *Model:* Although a sensitivity analysis suggested the clustering results were insensitive to the model, a larger cohort with more cases would allow a more complex statistical model, or the inclusion of more risk factors. Although the application of clustering methodologies to epidemiological data is becoming popular, methods to objectively determine the optimum number of clusters for a particular application have yet to be established. Most importantly, we found that disease identification and clustering was sensitive to the number of diseases, that in turn was surprisingly sensitive to the fitted model through the multiple-testing adjustments used to determine which diseases to include. *Causal associations:* We aimed to explore associations between diseases, but further work is needed to determine if the observed associations are causal.

## Strengths of methodology

Diseases were assessed and selected prior to the study, on the basis of clinical and epidemiological criteria. Established and interpretable statistical methodologies were used in new but statistically rigorous ways. Risk associations were calculated before clustering, providing advantages in terms of modelling and interpretation of results. Proportional hazards methods provided access to several decades of epidemiological experience, and are familiar to the medical community. Analyses were sex-specific, used (left-truncated) age as a time variable and multiple-adjustment to reduce the influence of correlations between risk factors, age in particular, and were censored by the first occurrence of cancer (other than non-melanoma skin cancers). Estimates were adjusted for likely confounders, and multiple adjustment will reduce the influence of correlations between risk factors on subsequent clustering. The resulting estimates are normally distributed, allowing rigorous (multivariate) statistical tests to compare the equivalence of risk factors for different diseases, and their marginal distributions are easy to calculate. This allowed adjustment for many known risk factors but to subsequently focus on a subset of the most biologically relevant factors by using marginalisation ^32^ to remove parameters of lesser interest. The procedure also ensured that each risk factor was represented by a single variable when clustering, avoiding clustering being dominated by e.g. a categorical variable with many different categories. Rigorous statistical tests were used to compare different diseases’ risk factors, clustering results were consistent with statistical tests, were relatively insensitive to changes in the proportional hazards model, and sensitivity analyses found no evidence that clustering was driven by prior disease. Distances between fits used estimated parameters and their covariance matrices, retaining as much information from the data as possible. Hierarchical clustering is easily visualised, and may help inform hierarchical disease classifications. Diseases were confirmed to cluster into clinically meaningful groups.

### Summary

The associations of common risk factors with disease incidence were used to characterise over 400 diseases in men and women, and to identify clusters of 78 diseases that were present in both sexes with statistically significant risk factors after a Bonferroni multiple-testing adjustment. We aimed to incorporate as much clinical and epidemiological knowledge as possible, and to adopt analyses that are easily interpretable, familiar to the medical community, and underpinned by a rigorous statistical methodology. The broad perspective gained from the simultaneous study of several hundred diseases emphasises that BMI can have a quantitatively different influence on disease risk for men and women, and that proportional hazards models are more likely to fail with more cases. Both of these important points should be considered in relevant epidemiological studies. We found that the associations of common risk factors with disease incidence was sufficiently specific to identify the equivalent disease in the opposite sex for 38% of 172 diseases studied here, and 80% have their opposite-sex pair among the nearest 8 diseases, suggesting that quantitatively similar risk factors may indicate similar underlying disease. This hypothesis was supported by hierarchical clustering, that tended to produce clinically similar clusters of diseases, and suggested several plausible but presently unconfirmed associations between disease. Some patterns of multimorbidity, such as a cluster of diseases linked to renal failure, are likely to be driven by common disease pathways and risk factors. All the diseases studied here are common causes of hospital admission, representing a substantial burden of ill health. We highlighted several symptoms of unknown causes (ICD-10 R-coded diseases), that appear to be linked with more clearly diagnosed disease, and emphasised the potential for hospital admissions to be biased by known risk factors for disease.

Overall, we have developed a methodology and demonstrated a proof of principle for clustering diseases in terms of their associations with established and easily measured risk factors. Future work is intended to optimise the approach, benchmark it in different datasets, and explore applications in diagnosis, prognosis, aetiological understanding, and drug development.

## Methods

### Data sources

UK Biobank data can be accessed by application through www.ukbiobank.ac.uk, along with relevant code and the disease selection dataset.

### Coding and diagnosis of disease

The National Clinical Coding Standards ^43^ define the primary diagnosis as the main symptom or disease treated, and arguably this primary cause of hospital admission provides the most reliable diagnosis. Additional diagnoses made after admission to hospital can correspond to less severe complaints diagnosed by chance, or occurring in association with either the primary or a different disease. Coding standards require that only diseases that affect the patient’s management should be recorded ^43^, which will not necessarily include all existing diseases. They are also biased by medical practice, with diagnoses limited to those that are investigated. Therefore the present study is restricted to the smaller number of primary diagnoses that were expected to have passed a threshold of severity, and were more likely to be unrelated to undiagnosed or co-occurring disease.

### Clinical considerations

Not all diagnosed and coded diseases are suitable for study. For example, a disease may have an uncertain diagnosis, or be unrelated to age or environmental exposures. Primarily we required that 3-digit ICD codes refer to a clear diagnosis of an age-related disease. Random events including accidents or infections due to a chance exposure were excluded unless modified by an underlying, possibly age-related, condition or predisposition. For example, some infectious diseases are more strongly influenced by chance lifestyle exposures than by age-related risks, but urinary tract and chest infections are influenced more by a weakened immune system than from a chance exposure alone, and were included. Diseases common before the start of the UK Biobank study such as pregnancy-related diseases were excluded due to insufficient cases. Any of the above, or related issues, can cause statistical models to fail or lose power, and we also excluded any diseases that failed any statistical test described later.

The above considerations led us to firstly exclude ICD-10 coded diseases beginning with: Z (factors influencing health status) - because not disease specific, Q (congenital) and O, P (Diseases related to pregnancy and perinatal period), U (new and antibiotic resistant diseases), V, X, Y (external causes of morbidity and mortality), and T (multiple injuries, burns, and poisoning) - usually reflecting a chance exposure. An epidemiology-trained pathologist (KG) selected and categorised diseases as excluded, acute-onset, chronic, due to infection, due to injury, or unknown aetiology (R-coded diseases in ICD-10, retained to allow follow-up studies).

### Selection at the 4-digit ICD-10 code level

Incidence data may be more informative if a 3-digit ICD-10 coded disease, is split into 4-digit coded disease subtypes. If these more accurately reflect the underlying aetiology, then associations with risk factors are expected to be clearer (with for an equivalent number of cases, smaller confidence intervals and bigger effect sizes). Therefore the 3-digit selections were examined and revised by a physician with training in epidemiology (IT). Where substantial aetiopathological differences existed, 3-digit codings were split into smaller groups. Often one or more 4-digit codes were excluded from a 3-digit group for a reason listed previously. Occasionally, diseases were split into a combination of one or more 4-digit codes and a grouping of 4-digit codes (see Supporting Information). The 4-digit selection was reviewed and tested for self-consistency to prevent typographical input errors. The selection is listed in the Supporting Information, table 1.

### Survival analysis

The survival analysis used a proportional hazards model ^44,45^ with age as the time variable, and the data were left-truncated at the age when participants attended the UK Biobank assessment centre. The data were right-censored if the end of the study period occurred before the disease of interest, or if there was any cancer other than non-melanoma skin cancer, because many cancers and cancer treatments are known to influence subsequent disease risk. Using age as the time variable allows strong age-dependencies to be accurately modelled through the baseline hazard. All calculations used R version 4.0.0.

The numerical measures of height, BMI, and SBP were standardised using their joint mean and standard deviation across men and women, smoking status was: never, previous, or current, alcohol consumption was: rarely (less than 3 times per month), sometimes (less than 3 times a week, but more than 3 per month), regularly (3 or more times each week), walking pace was: slow, average, brisk, and education was: degree level, post-16 (but below degree), to age 16 or unspecified. Baseline was taken as: no diabetes, never smoker, rarely drink, brisk walking pace, degree-level education, minimum deprivation tertile, and women with no children or HRT use. Analyses were multiply adjusted to minimise the influence of correlations between risk factors and capture as much causal information in the fitted parameters as possible. The measured risk factors had less than 1% missing values, allowing a complete case analysis. Because the risk factors are commonly measured, equivalent analyses in other datasets are possible. Sensitivity analyses with sex-dependent tertiles found similar results to those of the main text; see Supporting Information, figures 2 and 4.

### Statistical inclusion criteria

There is no general rule to determine how many cases are sufficient to ensure meaningful estimates for parameters and their covariances ^46^. We excluded diseases if their parameters or covariance matrices were undefined, or their covariance matrices’ eigenvalues were unusually large, indicating excessively large confidence intervals for one or more parameter (see Supporting Information, figure 1). This was typically due to insufficient data in one or more category, and were usually diseases that occur at the older (or younger) extremes of age range (e.g. delirium or excessive menstruation respectively), with too few cases in the younger (or older) YOB tertiles. To select a smaller set of diseases that have the most statistically significant risk factors and are easier to study and discuss, we excluded diseases whose risk factors were not statistically significant after a Bonferroni multiple-testing adjustment of a multivariate *χ*^2^ test for statistical significance of the fitted parameters. Finally the proportional hazards assumption was tested using a global *χ*^2^ test of the Schoenfeld residuals ^45^, and diseases failing the test after an FDR multiple-testing adjustment ^32^ were excluded. When testing for failure (and exclusion), an FDR adjustment is stricter than a Bonferroni adjustment and will exclude more diseases.

### Multivariate statistical tests and clustering metrics

Because maximum likelihood estimates for parameters e.g. 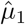 and 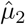 are approximately normally distributed, statistical tests are easy to construct. For 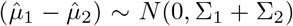 and 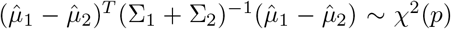, if they have the same mean with *µ*_1_ = *µ*_2_, then 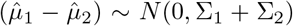 and 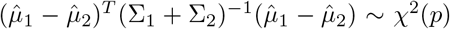 where *p* is the number of parameters ^46^. This was used to test the null hypothesis that the fitted parameters of diseases in men and women are the same, using the MLE estimates for the covariance matrices (figures 2 and 3). We also tested the null hypothesis that diseases in the same cluster have the same mean, by noting that,

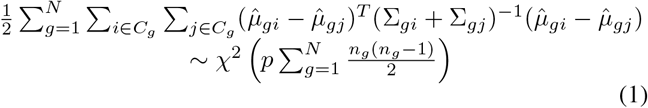

where *n*_*g*_ is the number of diseases in cluster *g* with members *C*_*g*_, *N* is the number of groups, and *p* is the number of fitting parameters. After removing the 8 diseases in figure 3 with statistically significant differences between men and women at the 0.05 level after an FDR multiple-testing adjustment ^32^, we plotted the left-hand side of Eq. 1 versus *N* to determine a minimum value of *N* = 63 where there is no longer a statistically significant difference at the 0.05 level (figure 5). However our main interest is in the similarity between risk factors for diseases, not whether they are statistically different. The left hand side of Eq. 1 falls rapidly until *N* ≃ 24, suggesting that most of the variation is captured in the first 24 clusters. This “elbow criterion” ^47^, was used in figures 4 and 5. Presently there is no rigorous and established method to determine how many clusters there should be ^46^.

**Figure 5:**
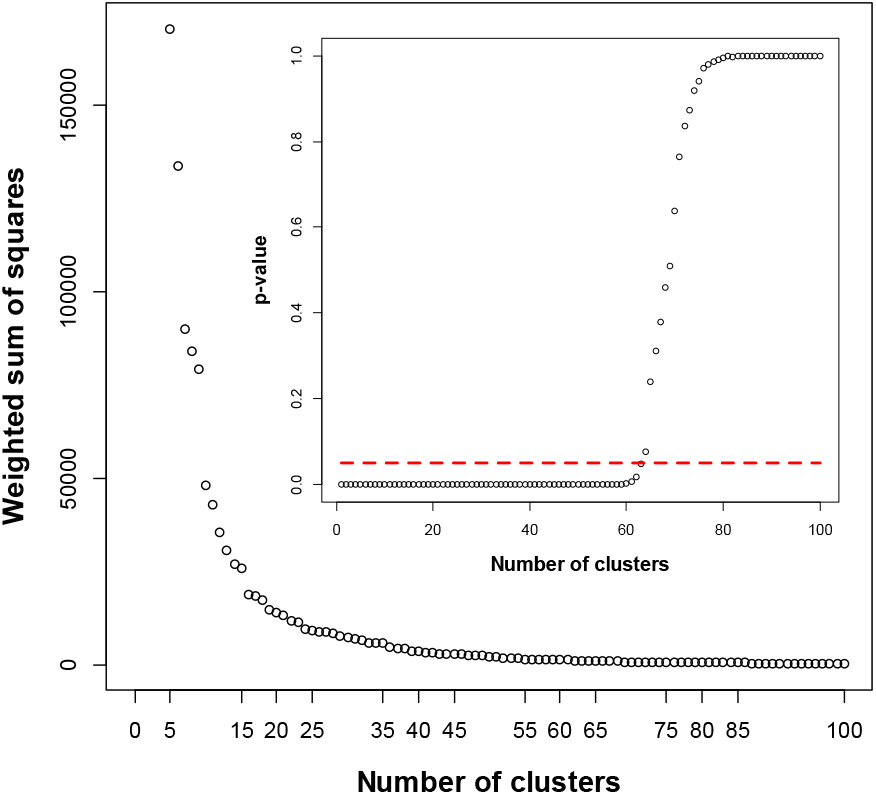
The “elbow” in the weighted sum of squares of differences in the fitted parameters in each cluster (Eq. 1), at ≃ 24 clusters, qualitatively indicates how many clusters to keep. With 63 or more clusters there are no statistically significant differences at the 0.05 level between fitted parameters in each cluster (inset).

A distance between fitted parameters must reflect both their value and their covariances, so that distances between poorly fitted parameters should be smaller than the distances between the same estimates with smaller covariances. This is true of the Battacharyya distance *D*_*B*_, that measures the similarity between probability distributions, and gives the difference between two multivariate normal distributions as,

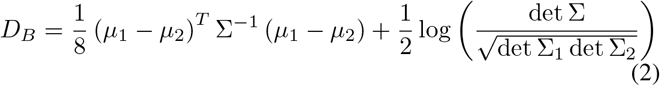

with Σ = (Σ_1_ + Σ_2_)*/*2. The first term is proportional to the *χ*^2^(*p*) that was used to test the null hypothesis of equal means (*µ*_1_ = *µ*_2_) in figures 2 and 3. As a consequence the largest p-values will tend to coincide with the smallest Battacharyya distances, but *D*_*B*_ also incorporates extra information from the estimated covariance matrices to compare the shape of the probability distributions. The minimum *D*_*B*_ can be used to assign a partner to each disease (figure 2). We hierarchically clustered the 156 diseases using *D*_*B*_ and the ward.D2 algorithm in the R software package. Diseases were assigned to 24 clusters, as suggested by the elbow criteria ^47^ and figure 5. The clustering is shown in figure 4, along with a heat map for the coefficients of each risk factor associated with each disease mapped onto a 0-1 scale using an inverse logit function.

### Sensitivity analysis

Meaningful disease clusterings must be insensitive to small changes in the data or to the model used to analyse it. In addition to visual comparisons, we quantified the differences between two clusterings of disease by considering the pairs of diseases that remain in the same cluster, independent of the clustering algorithm. Specifically, consider clustering the same set of diseases into e.g. 24 groups by two different algorithms *A* and *B*, such as using coefficients estimated from two different proportional hazards models. Take the observed number of all possible disease pairs within clusters in *A* as *n*_*A*_, the equivalent number in *B* as *n*_*B*_, and the number common to both as *n*_*AB*_. The maximum proportion of disease pairs that are clustered together by both *A* and *B* is *p*_*AB*_ = *n*_*AB*_*/*min(*n*_*A*_, *n*_*B*_). In practice, the sensitivity analyses produced clusterings with more disease pairs, and in this paper *p*_*AB*_ is the proportion of all clustered disease pairs that remain clustered together in the sensitivity analysis. Similar clusterings have *p*_*AB*_ *≃* 1, and unrelated clusterings have *p*_*AB*_ *≃* 0. Individual diseases that are particularly sensitive to the clustering procedures can be identified as those with no other diseases that are common to both their clusters (in *A* and *B*).

## Data Availability

Data are available from UK Biobank at www.ukbiobank.ac.uk, by application.

## Acknowledgements

This research has been conducted using the UK Biobank resource under application number 42583. Anthony Webster, Benjamin Cairns, and Iain Turnbull are supported by intermediate, senior, and clinical research fellowships respectively, from the Nuffield Department of Population Health (NDPH). Kezia Gaitskell is supported by a clinical lectureship from the National Institute for Health Research (NIHR, grant no. CL-2017-13-001). Robert Clarke is supported through core funding for NDPH from the Medical Research Council (MRC) and the British Heart Foundation (BHF). The University of Oxford MRC Population Health Research Unit is funded through a strategic partnership between the Medical Research Council and the University of Oxford. AJW thanks Katie E. Webster for helpful comments and suggestions.

## Author contributions

The project, methodology, and statistical analyses were developed by AJW, with comment from BJC and RC. Results were discussed with all authors. KG and IT selected 3-digit and 4-digit ICD-10 coded diseases respectively, with consistency checks and feedback from AJW. AJW wrote the paper, with comments from all authors, and extra references from KG and RC.

